# Mendelian randomization to inform a clinical trial of chitotriosidase inhibition for pulmonary sarcoidosis

**DOI:** 10.1101/2025.09.25.25336344

**Authors:** Daniel A. Culver, Marco F. Schmidt, Marlies S. Wijsenbeek-Lourens, Michael Kreuter, Vincent Cottin, Jolanta Kunikowska, Natalia V. Rivera, Marcel Veltkamp, Hannes A. Baukmann, Justin L. Cope, Charles N. J. Ravarani, Zbigniew Zasłona, Theodoros Charitos, Piotr S. Iwanowski

## Abstract

**Background:** Current sarcoidosis treatment is limited to glucocorticoids and immunosuppressants that provide symptomatic relief but carry significant side effects. Chitotriosidase 1 (CHIT1) has been correlated with disease severity and is highly expressed in sarcoidosis granulomas, making it an attractive therapeutic target for disease modification.

**Research design and methods:** We performed Mendelian Randomization (MR) analysis using published genome-wide association studies to evaluate the causal relationship between CHIT1 levels and sarcoidosis risk. Eight single nucleotide polymorphisms were selected as instrumental variables from multiple sarcoidosis datasets.

**Results:** MR demonstrated a significant positive causal effect of CHIT1 levels on sarcoidosis risk (p = 0.0355), reproduced across multiple datasets. Building on this genetic evidence, a proof-of-concept study with a CHIT1 inhibitor, OATD-01, was designed (KITE study) that represents an innovative approach highlighting academic-industry partnership to identify therapeutic targets, develop supporting data to de-risk clinical trial development, and implement novel trial endpoints to assess treatment promise. An innovative 18F-FDG-PET/CT-based primary endpoint was proposed for the KITE study to capture anti-granulomatous effects.

**Conclusions:** These genetic analyses support CHIT1 as a causal factor in sarcoidosis pathogenesis, providing strong rationale for targeting CHIT1 therapeutically. The evidence supports the ongoing development of OATD-01, a first-in-class CHIT1 inhibitor, currently being evaluated in the KITE study using this novel imaging biomarker approach that may provide a template for future sarcoidosis clinical trials.

**Trial Registration:** The trial is registered at ClinicalTrials.gov (CT.gov identifier: NCT06205121).

## Introduction

Sarcoidosis is a multisystem inflammatory disorder characterized by non-caseating granulomas, with pulmonary involvement occurring in 90% of cases. Current treatment options are limited to glucocorticoids and immunosuppressants, which provide symptomatic relief but might be associated with significant side effects [1]. Lack of approved disease-modifying therapies and more tolerable disease suppressing therapies represent unmet medical needs in sarcoidosis management [2]. The desired disease modification would entail preventing or reversing granuloma formation, halting progression to fibrosis, and achieving sustained remission without continuous immunosuppression. Despite extensive research efforts and the identification of various molecular targets including cytokines such as tumor necrosis factor (TNF), interleukin 6 (IL-6), chemokines, and specific immune cell populations, no disease-modifying therapy has been approved to date. This gap in therapeutic options particularly affects patients with chronic progressive disease who face significant morbidity and reduced quality of life, not infrequently further complicated by the side effects of currently available therapies Novel therapies are therefore required both on efficacy and tolerability/safety grounds.

Development of new treatment approaches has been limited by the absence of a clear regulatory pathway, perception that sarcoidosis is generally a non-bothersome, self-limiting disease, and insufficient understanding of disease pathobiology. Several preclinical granuloma models have been developed and employed over the past decade—these have directly facilitated pilot clinical studies as well as larger scale clinical trials. Use of these models, as well as hypothesis-generating unbiased approaches like genomic and proteomic assessment of human sarcoidosis samples has also led to identification of new molecular targets. These candidate therapeutic targets may be identified by single investigators, collaborative multi-institutional research groups, or biotechnology companies, but the pathway from identification to mechanistic evaluation to assessment in human sarcoidosis and finally clinical trials usually requires collaboration among all these stakeholders. Moreover, with a limited number of appropriate patients for explanatory trials, absence of validated surrogate endpoints and few expert centers, establishing confident links between candidate novel therapeutic targets and potential clinical utility is important to rationalize investment of time and financial resources. In this manuscript,, we aim to demonstrate the value of industry-academic collaboration by highlighting an example of how this partnership informed preclinical and design phases of an ongoing sarcoidosis trial (NCT06205121).

Circulating proteins (biomarker candidates) associated with granulomatous activity, disease severity, or future outcomes are attractive therapeutic candidates for drug development. One such biomarker, chitotriosidase 1 (CHIT1) has been correlated with disease severity, need for treatment and risk of deterioration [3,4]. It appears to be more specific than conventional biomarkers such as angiotensin-converting enzyme [5,6]. CHIT1 is highly expressed in sarcoidosis granulomas [7,8]. During monocyte-to-macrophage differentiation and subsequent macrophage activation, exuberant CHIT1 expression contributes to the inflammatory cascade, granuloma formation, and tissue remodeling that is characteristic of sarcoidosis. The pathologically activated macrophages, through pathways influenced by CHIT1, stimulate granuloma formation and fibroblast proliferation and further differentiation into myofibroblasts that may ultimately promote fibrosis [9]. Targeting CHIT1 could potentially interrupt both granuloma formation and subsequent fibrotic progression, addressing two hallmarks of sarcoidosis.

Recent observations suggest that CHIT1 inhibition might provide a novel disease-modifying approach for sarcoidosis treatment [8,10,11]. Immunocytochemical analysis of biopsies obtained from bronchoalveolar lavage fluid smears, bronchial mucosa, and mediastinal lymph nodes of patients with sarcoidosis demonstrates high expression of CHIT1, predominantly localized in granulomatous epithelioid and giant cells [8,12]. Uninvolved organs did not exhibit CHIT1 in macrophages. CHIT1 activity also significantly correlates with uptake of ^18^F-fluorodeoxyglucose ([^18^F] FDG) in positron emission tomography/computed tomography (PET/CT) imaging in patients with sarcoidosis, suggesting association with granulomatous activity [4].

## Methods

To further evaluate the strength of the relationship between CHIT1 and sarcoidosis, we performed Mendelian randomization (MR) analysis from published genome wide association studies (GWAS). The GWAS dataset for CHIT1 levels was retrieved from Guðjónsson *et al*. [13]. GWAS datasets for sarcoidosis were utilized from six sources: Sakaue *et al*. (1,938 cases, 662,622 controls) [14], the FinnGen research project (4,399 cases, 405,620 controls) [15], Fischer *et al*. (1,726 cases, 5,482 controls) [16], and three from the Neale Lab’s analysis of UK Biobank phenotypes [17]: self-reported sarcoidosis cases (705 cases, 360,436 controls), sarcoidosis diagnosed by a medical professional (395 cases, 91,392 controls), and sarcoidosis cases based on ICD-10 code D86 records (170 cases, 361,024 controls). Next, datasets were pre-processed with version 1.7.8 of the tool MungeSumstats [18], using default parameters that include checking to ensure variants are on the respective human reference genome (GRCh38 or GRCh37 depending on the datasets), removing duplicate SNPs, and removing multi-allelic SNPs (according to the SNP database db144).

First, pre-processed summary statistics files were read and thresholds were applied to the exposure SNPs to ensure they were valid instruments. The *p-*value threshold was *p* < 5E−08, as is standard for genome-wide experiments. Independent SNPs were identified by performing linkage disequilibrium (LD) clumping to discard SNPs in LD with another variant with a smaller *p*-value association based on the European reference panel from the 1000 Genomes Project [19] using the ld_clump() function of the R package ieugwasr [20], which provides a wrapper around PLINK [21]. LD clumping parameters used for this approach were a correlation r^2^ of 0.001 for variants within each haplotype block 10,000 kbp in size, which is the default used by the ld_clump() function.

If at least three SNPs remained as instrumental variables, the MR-Rücker framework [22] was applied to determine whether the inverse-variance weighted (IVW) [23] or the MR-Egger [24] method was best supported by the data. In addition, the MR-PRESSO test, which evaluates and corrects for horizontal pleiotropy via detection of outlier instrumental variables, and the weighted-median method [25], which is robust to outliers, were applied. If only two SNPs remained as instrumental variables, the IVW method was applied, or if only one, the Wald ratio method [26]. All methods were implemented in the TwoSampleMR package [22]. An MR experiment using a binary outcome produces an effect estimate, which corresponds to the log odds of the outcome per copy of effect allele [27], together with standard errors, from which 95% confidence intervals can be derived.

## Results

Based on the above methods, we selected eight single nucleotide polymorphisms (SNPs) as instrumental variables (SI Table 1). In this dataset, the estimated causal effect of CHIT1 levels on sarcoidosis was significant and positive (estimate = 0.0745, standard error = 0.0354, p = 0.0355; Figure 1), suggesting that higher CHIT1 levels might significantly increase the risk of sarcoidosis. This finding was supported by the robust weighted median MR method (estimate = 0.093, standard error = 0.0456, p = 0.0412; SI Figure 1). The MR-PRESSO global test was insignificant, meaning that horizontal pleitropy was not detected (RSSobs = 10.71057, p = 0.52) [28]. As no outliers were identified, the MR-PRESSO outlier test and distortion tests could not be applied.

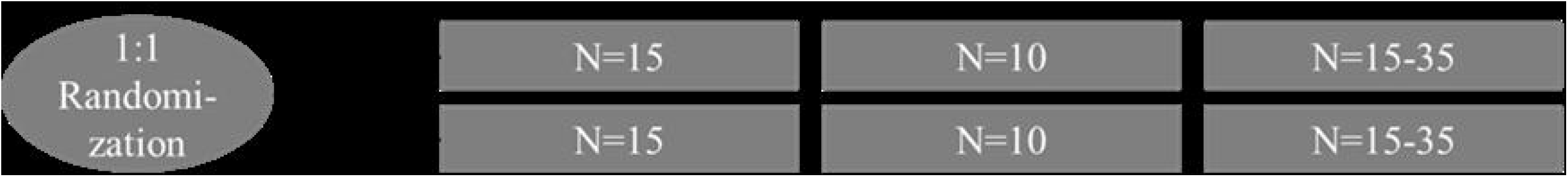

**Table 1.** Primary endpoint used in KITE study (ongoing)

| Target lesions, pulmonary | Target lesions, any location | Any new lesions | Overall response |
| --- | --- | --- | --- |
| No uptake above the background | No >25% increase of mean of $SUV_{max}$ from baseline | Absence | **Complete response** |
| Uptake above the background<br>AND<br>>25% decrease of mean of $SUV_{max}$ from baseline | No >25% increase of mean of $SUV_{max}$ from baseline | Absence | **Partial response** |
| Uptake above the background<br>AND<br>Mean of $SUV_{max} \leq 25\%$ decreased OR $\leq 25\%$ increased from baseline | No >25% increase of mean of $SUV_{max}$ from baseline | $\leq 2$ new lesions | **Stable metabolic disease** |
| Uptake above the background<br>AND<br>Mean of $SUV_{max} > 25\%$ increased from baseline | | $\geq 2$ new lesions | **Disease metabolic progression** |
| No evaluable data and no otherwise evidence of disease metabolic progression |  |  | **Indeterminate response** |

SUV = Standardized Uptake Value

These results of our primary analysis were further investigated in a reproduction analysis with four additional datasets for sarcoidosis, but we were unable to utilize the Fischer GWAS as the relevant SNPs were not included in it. The reported results were supported by separate analysis of GWAS data from both Finnish and one of the United Kingdom populations [15,17]. Two of the UK estimates (for which the number of sarcoidosis cases in the source data was quite small) did not reach statistical significance (SI Figure 2). Together, these results established a more solid evidence base impacting an independent role for CHIT1 in sarcoidosis pathogenesis, de-risking the decision to proceed with clinical trial development.

## Discussion

Extant genetic data suggest a role for CHIT1 in the pathogenesis of sarcoidosis. The current study lends support to the genetic analyses by using MR to demonstrate a relationship between CHIT1 levels and disease risk. In a reported cohort of 180 patients with sarcoidosis, those with homozygous *CHIT1* 24-base pair duplication (H/H genotype), resulting in complete absence of active CHIT1, showed better clinical outcomes at 3 years (p=0.025) and demonstrated a trend toward better outcomes at 5 years (p=0.090) [29]. Notably, H/H subjects had a significantly higher frequency of Löfgren syndrome, a phenotype associated with favorable prognosis and reduced macrophage activation [29]. Taken together, the current results and pre-existing genetic data align with the possibility that CHIT1 inhibition could attenuate mechanisms of granulomatous inflammation.

OATD-01 is a first-in-class small-molecule inhibitor of CHIT1 we identified as a candidate for sarcoidosis treatment [30]. *In vivo* efficacy of OATD-01 has been demonstrated in several models of inflammatory diseases such as chronic and acute asthma, inflammatory bowel disease, idiopathic pulmonary fibrosis, and non-alcoholic steatohepatitis [31,32,33]. OATD-01 has been evaluated in a murine model of granulomatous inflammation using multiwalled carbon nanotubules (MWCT) and ESAT6 [8,34]. When administered concomitantly with the MWCT, OATD-01 showed anti-inflammatory effects at the 10-day mark (acute model), reducing the percentage of neutrophils and proinflammatory cytokine CCL4 concentration in the epithelial lining fluid. In a parallel experiment where OATD-01 was not started until after granulomas were established (Day 10-chronic model) inhibition of CHIT1 led to a decrease in the number of organized lung granulomas and the expression of sarcoidosis-associated genes [8].

In a short-term clinical study, such as a proof-of-concept trial, conventional respiratory function markers like forced vital capacity are unlikely to be sensitive to detect biologic effect. Novel surrogate markers are appealing in this setting, and these should directly evaluate inflammatory burden. Inflammation in macrophages depends on the process of glycolysis. The latter can be measured and imaged by fluorine-18 fluorodexyglucose positron emission tomography with CT attenuation correction ([^18^F]FDG PET/CT). In pulmonary sarcoidosis, evidence of PET/CT uptake changes correlating with changes in biochemical biomarkers of T-cell activation and macrophage-derived granulomatous inflammation suggest that [^18^F]FDG PET/CT is a good imaging biomarker for assessment of inflammation as there is no direct relation between glycolysis and these biochemical biomarkers [35]. Genetic deficiency or pharmacological inhibition of CHIT1 affects cellular metabolism, by inhibiting glycolysis resulting in an anti-inflammatory macrophage phenotype. Specifically, macrophages treated with OATD-01 have attenuated glucose uptake, which impedes glycolysis and subsequent inflammation [36]. It was demonstrated that the [^18^F]FDG PET/CT maximum standardized uptake value (SUV_max_) and CHIT1 plasma activity significantly correlate in patients with sarcoidosis regardless of gender, age, duration of disease and radiography stage (as opposed to angiotensin-converting enzyme, another sarcoidosis biomarker) [37]. These data support utilizing [^18^F]FDG PET/CT as the imaging technique to capture antigranulomatous/anti-inflammatory effect of OATD-01 in pulmonary sarcoidosis, taking into account that this technique is otherwise already useful for the diagnosis of this condition [38].

Although [^18^F]FDG PET/CT may be a useful read-out of biologic activity from an intervention such as OATD-01, a positive correlation would not necessarily validate [^18^F]FDG parameters for use in clinical practice [39]. At present, [^18^F]FDG uptake does not satisfy the requirements for a valid clinically relevant surrogate endpoint in pulmonary sarcoidosis. However, a recent meta-analysis of studies evaluating the prognostic value of [^18^F]FDG PET/CT (11 studies, n=308) demonstrated its good correlation with functional scores (i.e., reduction in SUV_max_ correlated significantly with FVC improvement) [40]. All in all, [^18^F]FDG PET/CT has emerged as an attractive outcome measure and a proof-of-concept marker of biologic effect from an intervention, especially when a clinical trial is short enough to avoid a high likelihood of spontaneous resolution. In fact, [^18^F]FDG PET/CT has been utilized in a few academic clinical studies evaluating response to potential therapies for pulmonary or multiorgan sarcoidosis, specifically glucocorticoids [41,42], anti-TNF therapies [43,44], or tofacitinib [45]. Some methodology variations have also been tested with a volumetric modality for analysis of global lung inflammation [46] or quantifying glycolysis of the entire lung [47] in parallel to the standard evaluation of the SUV_max_. Thus, [^18^F]FDG PET/CT-based efficacy measure has been adopted in clinical trials of novel compounds over the last few years, though as secondary endpoints [NCT02888080, NCT04064242]. The utility of various endpoints for pulmonary sarcoidosis trials remains to be an emerging topic under discussion by the sarcoidosis clinical research societies [48,49].

Taking into account preceding research and observations, a primary endpoint based on [^18^F]FDG PET/CT has been proposed for the proof-of-concept clinical study with OATD-01 (the KITE study; see below). Unlike most earlier adoptions of [^18^F]FDG PET/CT technique for measurement of treatment efficacy based on plain SUV measurements (SUV_max_ in particular), the KITE study is pursuing a semi-qualitative analysis adopting the approach developed by the European Organization for Research and Treatment of Cancer (EORTC) for [^18^F]FDG PET/CT-based evaluation of efficacy in oncology trials [50]. This includes a quantitative evaluation of the target lesions, focused on the pulmonary target lesions (up to 5 per location), but taking into account also the image changes in the mediastinal and extrathoracic locations (up to 5) as well as appearance of non-target lesions post-baseline, if any. The response to study treatment is assessed as complete at end of treatment in case of no uptake above the background in pulmonary target lesions with no new lesions in any location and no increase >25% of the mean SUV_max_ in target lesions in mediastinal and extrathoracic locations. A new lesion denotes one which is [^18^F]FDG avid with an uptake greater than twice that of the surrounding tissue on the attenuation corrected image. The empirical 25% cut-off point for the ‘stable’ image has been proposed, adopted from the current medical practice when evaluating [^18^F]FDG PET/CT (glucose metabolism) changes in sarcoidosis as well as the EORTC recommendations for tumor evaluations in clinical oncology [50], also taking into account the PET Response Criteria in Solid Tumors (PERCIST) [51]. Further response classification, depending on the uptake compared with the background in target and any new lesions, will declare partial response, stable metabolic disease, disease progression or indeterminate. The primary endpoint is presented in detail in Table 1.

This ongoing Phase 2 proof-of-concept study employs a randomized, double-blind, placebo-controlled, adaptive, multicenter design to evaluate OATD-01 in patients with active pulmonary sarcoidosis across approximately 20-30 sites in the United States and Europe [53]. Adult subjects (≥18 years) with symptomatic pulmonary sarcoidosis diagnosed according to criteria adopted from the American Thoracic Society guideline [38] with active granulomatous process demonstrated by [^1^□F]FDG PET/CT imaging are being randomized 1:1 to receive either OATD-01 25 mg or placebo orally once daily for 12 weeks. The patient profile corresponds to the low risk tier for morbidity or mortality as defined by the European Respiratory Society therapeutic guideline [54], including both treatment-naïve patients and those previously treated but currently untreated, with randomization stratified by prior treatment status (Figure 2). The total sample size ranges from 80-120 subjects (expected average: 96), with the final stage determined by conditional power calculations maintaining 90% power. Secondary efficacy endpoints include quantitative changes in [^1^□F]FDG PET/CT SUV parameters (SUV_max_, SUV_mean_, SUV_peak_) and lesion volumes across pulmonary parenchyma, mediastinal/hilar nodes, and extrathoracic locations, along with changes in pulmonary function (FVC, FEV□) and quality of life measures (Kings Sarcoidosis Questionnaire and Fatigue Assessment Scale). Additional secondary objectives encompass comprehensive safety and tolerability assessments, pharmacokinetic characterization, and pharmacodynamic evaluation of CHIT1 inhibition.

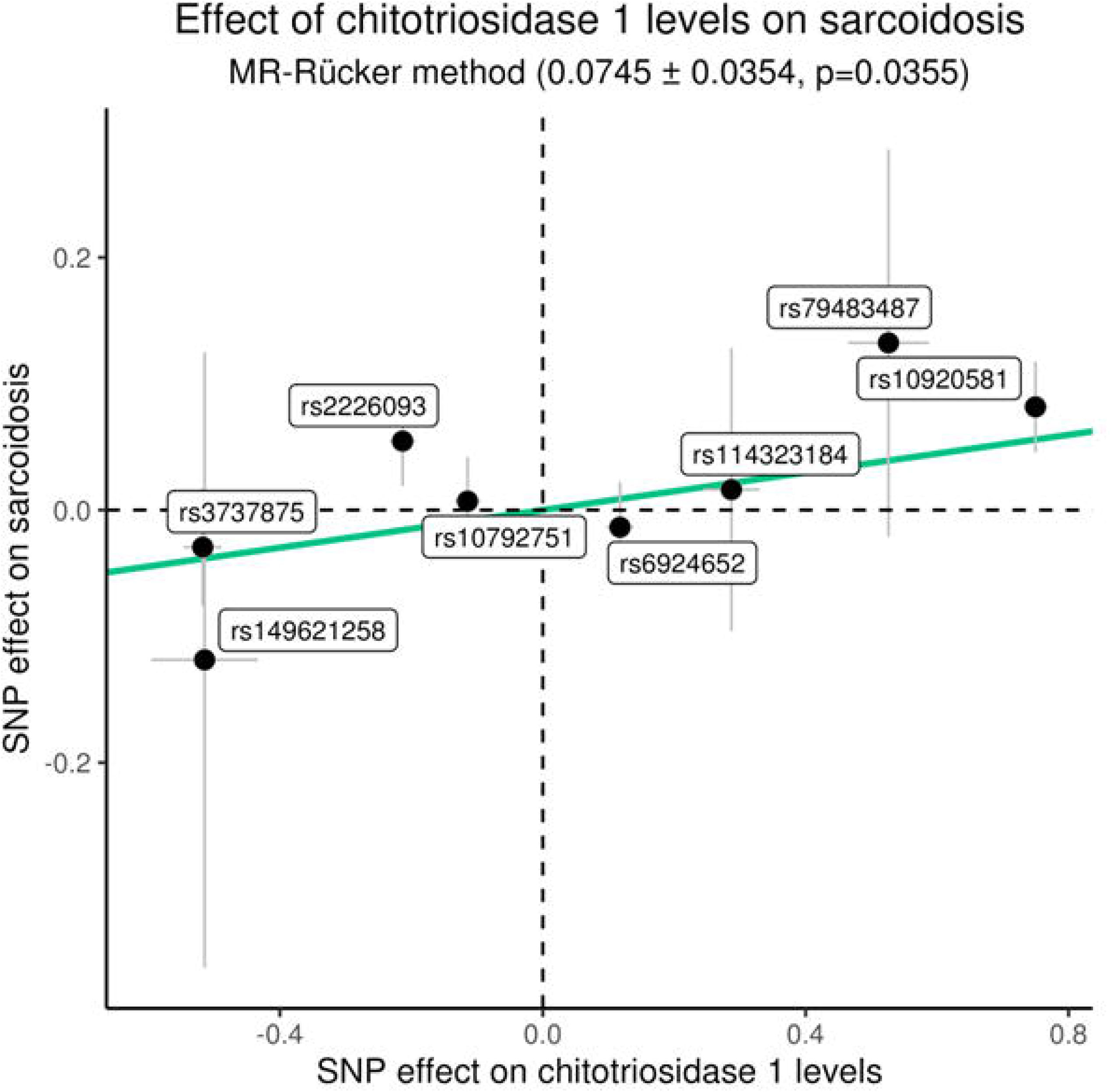

## Conclusions

This project demonstrates how industry-academic collaboration can lead to robust trial design in sarcoidosis. OATD-01 is a candidate in clinical development for pulmonary sarcoidosis, supported by multiple complementary lines of evidence. The scientific rationale for targeting CHIT1 is reinforced by several key findings: (1) mechanistic studies demonstrating the role of CHIT1 in granuloma formation and fibrotic progression, (2) genetic evidence from MR analyses showing a causal relationship between CHIT1 levels and sarcoidosis risk, (3) clinical observations linking CHIT1 polymorphisms to disease severity and outcome, and (4) preclinical data showing effectiveness of OATD-01 in relevant disease models.

The ongoing KITE study represents an innovative approach highlighting the partnership of academic and industry investigators to identify new therapeutic targets, develop additional supporting data to de-risk clinical trial development, and then implement novel and feasible assessments (trial endpoints) to more quickly assess whether the overall approach is promising. Surrogate markers of granulomatous inflammation, such as FDG uptake, complemented by conventional measures of organ function and quality of life may provide a template for future clinical trials in pulmonary sarcoidosis. This is particularly relevant given the current lack of regulatory guidance (e.g. by US Food and Drug Administration or European Medicines Agency) on optimal trial design and endpoints in this disease area.

The development of OATD-01 and other novel compounds for sarcoidosis, address an unmet medical need. Such a need for new therapeutic options has been concluded e.g. at the FDA Patient-Focused Drug Development meeting dedicated to sarcoidosis that took place in late 2024 [55]. The prospective KITE study results will not only inform further development of OATD-01 itself but may also advance the understanding of clinical trial methodology in pulmonary sarcoidosis, possibly contributing to future therapeutic development in this challenging disease area.

## Data Availability

All data produced in the present study are available upon reasonable request to the authors, subject to the clinical trial design outlined in this manuscript being ongoing at the time of submission of this manuscript

