## Supplementary material for "Mendelian randomization to inform a clinical trial of chitotriosidase inhibition for pulmonary sarcoidosis": SI Table 1

### Frequencies of effect alleles in exposure and outcome summary statistics

| **SNP** | **Chr** | **Position** | **EA** | **Exposure EAF** | **Outcome EAF** |
| --- | --- | --- | --- | --- | --- |
| rs114323184 | 1 | 202686551 | A | 0.05065 | 0.0333346 |
| rs79483487 | 1 | 203039108 | A | 0.02343 | 0.0149705 |
| rs3737875 | 1 | 203171543 | A | 0.1141 | 0.1778779 |
| rs10920581 | 1 | 203207271 | C | 0.2775 | 0.3164676 |
| rs149621258 | 1 | 203339473 | C | 0.01397 | 0.0095133 |
| rs2226093 | 1 | 203451086 | G | 0.3182 | 0.3229913 |
| rs6924652 | 6 | 154484559 | A | 0.2969 | 0.2981354 |
| rs10792751 | 11 | 84276862 | G | 0.6357 | 0.5830874 |

Chr = Chromosome, EA = Effect allele, EAF = Effect allele frequency; Exposure summary statistics are from Guðjónsson *et al*. [‎13] and outcome summary statistics are from Sakaue *et al*. [‎14].
