## Supplementary figures and images for "Mendelian randomization to inform a clinical trial of chitotriosidase inhibition for pulmonary sarcoidosis"

### SI Figure 1

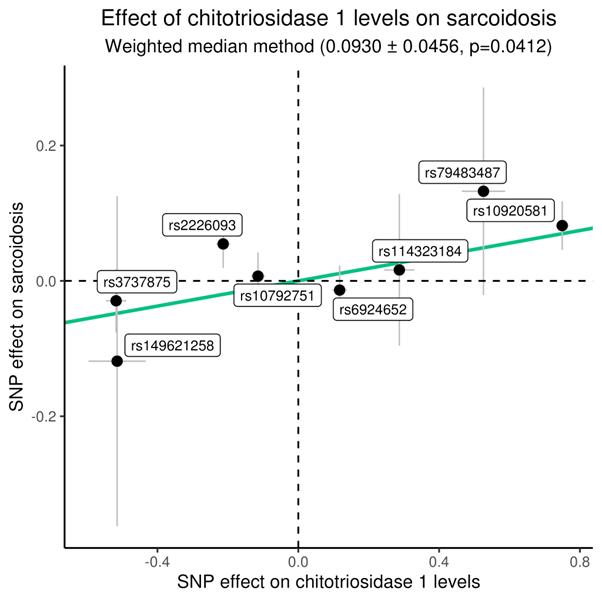

### SI Figure 2

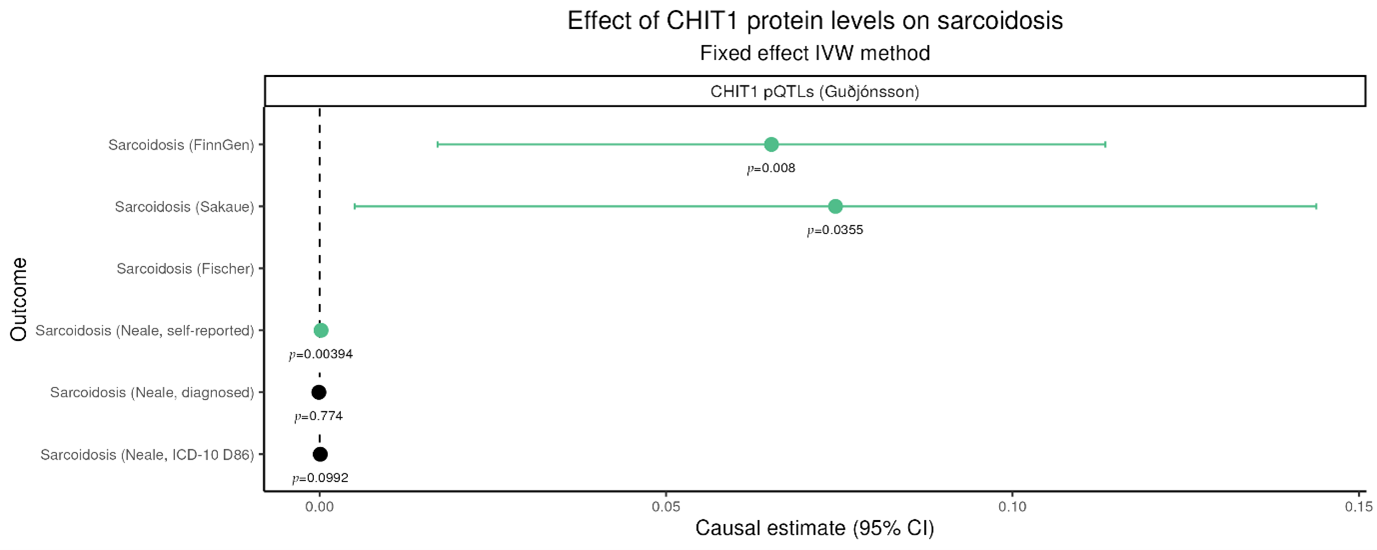
